# Patient satisfaction with telemedicine in the Philippines during the COVID-19 pandemic

**DOI:** 10.1101/2022.05.21.22274939

**Authors:** Alicia Victoria G. Noceda, Lianne Margot M. Acierto, Morvenn Chaimek C. Bertiz, David Emmanuel H. Dionisio, Chelsea Beatrice L. Laurito, Girrard Alphonse T. Sanchez, Arianna Maever L. Amit

## Abstract

**Introduction:** The capacity to deliver essential health services has been negatively impacted by the COVID-19 pandemic particularly due to lockdown restrictions. Telemedicine provides a safe, efficient, and effective solution that addresses the needs of patients and the health system. However, there remain implementation challenges and barriers to patient adoption in resource-limited settings as in the Philippines. This study thus aimed to describe patient perspectives and experiences with telemedicine services, and explore the factors that influence telemedicine use and satisfaction.

**Methods:** This study used a mixed-methods design through online surveys and in-depth interviews. An online survey using Consumer Assessment of Healthcare Providers and Systems (CAHPS) Clinician & Group Adult Visit Survey 4.0 (beta) and Telehealth Usability Questionnaire (TUQ) was accomplished by 200 participants aged 18 to 65 years. A subsample of 16 participants was interviewed to provide insights to the quantitative data. We used descriptive statistics to analyze survey data and grounded theory to analyze data from interviews.

**Results:** Participants were generally satisfied with telemedicine services, with most reporting that this was an efficient and convenient alternative to face-to-face consultations. However, only 2 in 5 perceived telemedicine as affordable. Our quantitative findings suggest that participants preferred telemedicine services rather than in-person consultations, especially in cases where they feel that their condition is not urgent and does not need extensive physical examination. Safety against COVID-19, and the availability of multiple communication platforms contributed to patient satisfaction with telemedicine. Negative perceptions of patients on their telemedicine provider, perceived higher costs, poor connectivity and other technological issues were found to be barriers to patient satisfaction.

**Discussion:** Telemedicine is viewed as a safe and efficient alternative to receiving care. Continued adoption of telemedicine will require improvements in technology and better patient communication related to their telemedicine provider and the associated costs.

## INTRODUCTION

The COVID-19 pandemic has negatively impacted the capacity to deliver essential health services especially among low-and-middle income countries such as the Philippines (1). Hospital admissions and procedures declined as lockdown restrictions were imposed in the country (2). Telemedicine provides an opportunity to minimize exposure to health workers and patients, while allowing patients to access high-quality healthcare that is safe, efficient, and cost-effective (3,4). Further, the utilization of telemedicine for non-urgent cases reduces the surge of outpatient visits following the COVID-19 crisis (5).

Although telemedicine has been recognized globally as a viable option to expand the reach of limited healthcare providers and resources during the pandemic, challenges including potential costs, limited technical resources, data privacy issues, and risks to patient safety pose problems to its wide-scale implementation (6,8). In the Philippines, health care and services were predominantly delivered through face-to-face means with an increase in the use of telemedicine especially in urban settings during the pandemic (7). Studying patient satisfaction is critical to guide action plans for the quality improvement of telemedicine services (8,9). To date, only three quantitative local studies have documented patient perspectives and experiences with telemedicine services during the pandemic (10–12). Our study builds on the existing evidence and aimed to provide a more in-depth insight into telemedicine use and satisfaction through the lens of patients in a low-an-middle income country. In better understanding patient experience and satisfaction with telemedicine, this study may provide insights into opportunities for integrating telemedicine into routine care and improving telemedicine services for widespread adoption even beyond the pandemic.

## METHODS

### Study Design

This study used an explanatory mixed-methods design consisting of an online survey and in-depth interviews. The qualitative component was guided by grounded theory to study concrete realities of participants and experiences using telemedicine services to render a conceptual understanding of patient satisfaction through an inductive, iterative, and interactive method (13).

### Study Participants

Participants were individuals aged 18 to 65 who reside in the Philippines and received telemedicine services during the COVID-19 pandemic.

### Sampling and Study Size

Convenience sampling was used given the logistical constraints to conduct field data collection during the pandemic. For the online survey, participants were invited through networks, telemedicine providers, Facebook, and Instagram. A subsample of the survey participants was invited for an in-depth interview. We selected them based on age, sex, location, and survey answers relating to their telemedicine experience and satisfaction to allow maximum variation sampling, which aims to capture as many population contexts as possible. The chosen respondents were individually contacted using contact details they provided in the survey through text or email. A total of 200 participants answered the online survey and 16 of them were interviewed.

### Instruments and Measures

The online survey questionnaire consisted of items on key socio-demographic characteristics and health-related expenditures, and questions from two validated instruments: 15 questions from Consumer Assessment of Healthcare Providers and Systems (CAHPS) Clinician & Group Adult Visit Survey 4.0 (beta) and 11 questions from Telehealth Usability Questionnaire (TUQ) (14,15). CAHPS is a registered trademark of the Agency for Healthcare Research and Quality (AHRQ) with the purpose of advancing our scientific understanding of patient experience with healthcare. TUQ was designed to be a comprehensive questionnaire that covers all usability factors, including usefulness, ease of use, effectiveness, reliability, and satisfaction. The TUQ has acceptable construct validity and internal consistency (16–18). Levels of patient satisfaction were measured for six components (convenience, communication, patient-physician relationship, cost, access, overall satisfaction) using a 5-point Likert scale to rate responses (1: strongly disagree; 2: disagree; 3: neutral; 4: disagree; 5: strongly agree). Participants were asked how they found out about telemedicine: advertising/paid promotions/endorsements, news, personal research, recommendations by friends or a health professional, social media, or through other means. Participants were also asked on the telemedicine platforms used: SMS (text message), messaging applications (e.g., Facebook Messenger, Viber, email, video call, voice call), telemedicine-specific platforms (e.g., KonsultaMD, SeeYouDoc, Aide mobile app, ClinicKo, Kitika, KonsultaMD, Medgate, SeeYouDoc, SeriousMD), and others not in the options. Comparisons of the quality of services delivered through telemedicine and in-person were measured using a 5-point Likert scale of agreement to the following statements: ‘Telemedicine services are the same as in-person consultations” and “Telemedicine services are better than in-person consultations”. A more in-depth response was obtained in the interviews, probing on their telemedicine use and experience, reasons for preferring or not preferring telemedicine over face-to-face, and the factors influencing their telemedicine use and satisfaction.

### Data Collection Procedures

We collected data through an online survey and online interviews from July to November 2021. We used Google Forms for the online survey, while Zoom and Google Meet were used for the interviews. We pre-tested the questionnaire and interview guide in English and Filipino among 15 participants who were similar in characteristics to our study population. The pre-test was conducted online in the same manner as a full-scale survey and assessed administration, organization, and content. The survey was improved based on the comments during the pre-testing phase. All survey participants were asked if they were interested in participating in the interview. Among those who consented, we invited participants for an interview through a video call platform (i.e., Zoom, Google Meet) chosen by the respondents. Each interview lasted anywhere from 30 to 120 minutes. Each interview participant was given approximately USD 3 (USD 1 = PhP 52 as of 11 May 2022) worth of token for participation. Interviews were conducted until data saturation was reached (19). All participants consented to the interview being recorded.

### Data Analysis

#### Quantitative Analysis

We analyzed our quantitative data using descriptive statistics: percentage for categorical variables, and median for continuous variables using SPSS Statistics version 25.0 (20). We described participants according to their age in years, sex (male or female), setting of residence (urban or rural), residence by island group (Luzon, Visayas, Mindanao), educational attainment (secondary or lower, college, post-graduate), employment status (full-time employment, part-time employment, unemployed, student, retired), monthly household income, monthly household health expenditure, and monthly individual health expenditure, membership to any health insurance (yes, no), overall health rating measured as a 5-point Likert scale. Levels of patient satisfaction were measured by computing the frequency and percentage for each item. This analysis is consistent with a study by Ackerman (21) that used TUQ to assess patients’ utilization of and satisfaction with telemental health in the perinatal period. For comparisons between telemedicine services and in-person consultations, we computed the frequency and percentage for both questions with respect to those who answered ‘agree’ and ‘strongly agree’. We classified those disagreed or strongly disagreed that telemedicine is better than in-person under the theme, “Telemedicine services are inferior to in-person visits”.

#### Qualitative Analysis

All interviews were audio-recorded, transcribed verbatim, and translated from Filipino to English. The researchers are native and/or fluent speakers of the two languages. Each participant was assigned a code to maintain anonymity. Inductive analysis was used to identify emergent themes and patterns from the qualitative data focusing on experiences and satisfaction with telemedicine services, guided by the principles of grounded theory (22). Transcripts of the interviews were read to identify themes and two research members independently coded the interviews according to themes. Each interview was coded according to general themes: facilitators or barriers to telemedicine use and satisfaction. Patterns from codes were used to further generate themes, which are central organizing concepts. The research team reviewed and finalized the themes. Any disagreements were resolved through a consensus. The quotes presented in this paper are either in the original English or translated from Filipino.

### Ethics Statement

This research was given ethical approval by the Ateneo University Research Ethics Committee (SMPH 2021 Group 15). Only those who consented to participate accomplished the online survey. Digital written informed consent was obtained from participants prior to the interview.

## RESULTS

### Participant Characteristics

Our survey participants had a median age of 31.50 years (IQR 23.5-46.0 years). More than half of our study population were female (64.0%) and college graduates (65.0%). A total of 62 participants worked full-time (31.0%) and 162 resided in Luzon (81.0%). The median monthly household income of participants was USD 577, median monthly household health expenditure was USD 69, and median monthly individual health expenditure was USD 38. Only half availed of any health insurance (51.5%). The median overall health rating of participants was 4 out of 5 (**Table 1**).

**Table 1.** Characteristics of survey respondents (n=200)

| Characteristic | N (%)<sup>a</sup> |
| --- | --- |
| Median age (in years (IQR)) | 31.50 (23.5 – 46.0) |
| Sex |  |
| Female | 128 (64.0) |
| Male | 72 (36.0) |
| Location by urbanicity |  |
| Urban | 170 (85.0) |
| Rural | 30 (15.0) |
| Residence by island group |  |
| Luzon | 162 (81.0) |
| Visayas | 28 (14.0) |
| Mindanao | 10 (5.0) |
| Highest Educational Attainment |  |
| Secondary or lower | 35 (17.5) |
| College | 130 (65.0) |
| Post-graduate | 35 (17.5) |
| Employment Status |  |
| Full-time employment | 62 (31.0) |
| Part-time employment | 15 (7.5) |
| Unemployed | 36 (18.0) |
| Student | 48 (24.0) |
| Retired | 8 (4.0) |
| Median Monthly Household Income in USD (IQR) | 577 (231 – 1,923) |
| Median Monthly Household Health Expenditure in USD (IQR) | 69 (38 – 115) |
| Median Monthly Individual Health Expenditure in USD (IQR) | 38 (19 – 77) |
| Avails Any Health Insurance |  |
| Yes | 103 (51.5) |
| No | 97 (48.5) |
| Median Overall Health Rating (IQR) | 4 (3 – 4) |

<sup>a</sup>May not total to 100% because of missing data

The median age in years of the 16 interview participants was 29. More than half (62.5%) were female. Thirteen (81.25%) participants were from Luzon, two (12.5%) were from Visayas, and one (6.25%) was from Mindanao. More than half (68.75%) of the participants availed of health insurance. Only seven (43.75%) participants disclosed their monthly household income with a median of USD 577.

### Overall Patient Satisfaction

Across all statements, most participants strongly agreed that they are satisfied with telemedicine services in terms of convenience, communication, patient-physician relationship, cost, and access (**Table 2**). Of these reasons, convenience was identified by majority of the participants (75.5%) to positively influence their satisfaction with telemedicine, saving them time from traveling to a hospital or specialist clinic. On the other hand, only 2 in 5 perceived telemedicine to be affordable. A total of 116 survey participants (58.0%) strongly agreed that they would use telemedicine services again.

**Table 2.** Levels of patient satisfaction with telemedicine services during the COVID-19 pandemic (n=200)

|  | Strongly Disagree<br>N (%) | Disagree<br>N (%) | Neutral<br>N (%) | Agree<br>N (%) | Strongly Agree<br>N (%) |
| --- | --- | --- | --- | --- | --- |
| **Convenience** |  |  |  |  |  |
| 1. The platform was easy to use. | 3 (1.5) | 2 (1.0) | 15 (7.5) | 72 (36.0) | 108 (54.0) |
| 2. Telemedicine saves me time traveling to a hospital or specialist clinic. | 0 (0.0) | 1 (0.5) | 17 (8.5) | 31 (15.5) | 151 (75.5) |
| **Communication** |  |  |  |  |  |
| 3. During my most recent visit, the healthcare provider and I were able to hear/communicate with each other clearly. | 0 (0.0) | 1 (0.5) | 25 (12.5) | 57 (28.5) | 117 (58.5) |
| 4. My most recent visit started on time. | 6 (3.0) | 17 (8.5) | 22 (11.0) | 62 (31.0) | 93 (46.5) |
| 5. Telemedicine provides for my healthcare needs. | 2 (1.0) | 13 (6.5) | 42 (21.0) | 57 (28.5) | 86 (43.0) |
| 6. Telemedicine is an acceptable way to receive healthcare services. | 3 (1.5) | 12 (6.0) | 42 (21.0) | 70 (35.0) | 73 (36.5) |
| 7. During my most recent visit, the provider explained things in a way that was easy to understand. | 0 (0.0) | 1 (0.5) | 11 (5.5) | 56 (28.0) | 132 (66.0) |
| 8. I feel comfortable communicating with the clinician using the telemedicine platform. | 2 (1.0) | 12 (6.0) | 38 (19.0) | 65 (32.5) | 83 (41.5) |
| **Patient-Physician Relationship** |  |  |  |  |  |

|  | **Strongly Disagree**<br>N (%) | **Disagree**<br>N (%) | **Neutral**<br>N (%) | **Agree**<br>N (%) | **Strongly Agree**<br>N (%) |
| --- | --- | --- | --- | --- | --- |
| 9. During my most recent visit, the provider listened carefully to me. | 1 (0.5) | 2 (1.0) | 12 (6.0) | 51 (25.5) | 134 (67.0) |
| 10. During my most recent visit, the provider showed respect for what I had to say. | 0 (0.0) | 3 (1.5) | 8 (4.0) | 48 (24.0) | 141 (70.5) |
| 11. During my most recent visit, the provider spent enough time with me. | 1 (0.5) | 1 (0.5) | 20 (10.0) | 58 (29.0) | 120 (60.0) |
| 12. During my most recent visit, the provider already had the medical information they needed about me. | 8 (4.0) | 8 (4.0) | 22 (11.0) | 52 (26.0) | 110 (55.0) |
| **Cost** |  |  |  |  |  |
| 13. Telemedicine is affordable. | 10 (5.0) | 15 (7.5) | 47 (23.5) | 50 (25.0) | 78 (39.0) |
| **Access** |  |  |  |  |  |
| 14. It is not difficult for me to avail telemedicine services. | 7 (3.5) | 9 (4.5) | 24 (12.0) | 55 (27.5) | 105 (52.5) |
| 15. My connectivity allows me to easily use telemedicine. | 0 (0.0) | 5 (2.5) | 28 (14.0) | 46 (23.0) | 121 (60.5) |
| **Overall Satisfaction** |  |  |  |  |  |
| 16. I would use telemedicine services again. | 0 (0.0) | 7 (3.5) | 31 (15.5) | 46 (23.0) | 116 (58.0) |
| 17. Overall, I am satisfied with my telemedicine experience. | 1 (0.5) | 6 (3.0) | 29 (14.5) | 65 (32.5) | 99 (49.5) |

### Comparisons between telemedicine and in-person consultations

#### Telemedicine services are the same as in-person consultations

A total of 88 survey participants agreed (44.0%) that the service provided through telemedicine was the same as in-person consultations (**Table 2**). This is supported by our qualitative findings that just like face-to-face consultations, telemedicine allows patients to access services provided by physicians, express their medical concerns, and have their concerns addressed. This perception of adequacy of care provided via telemedicine promotes its use:

> “[In a way,] telemedicine is the same [as face-to-face consultation] because I still get to talk to a doctor. You get to voice out your problems or your medical history, and then get a prescription or diagnosis.” (, 16-20, female)

#### Telemedicine services are better than in-person consultations

A total of 72 survey participants (36.0%) perceived telemedicine services to be better than in-person consultations. One participant who used KonsultaMD for a skin condition mentioned convenience and experiencing better quality of service:

> “[Telemedicine is] just so much more efficient and convenient, and I feel like the doctor’s not in a rush to get to the next patient, and they really try to [serve] you better over telehealth as compared to face to face consultations.” (21-25, female)

#### Telemedicine services are inferior to in-person visits

A total of 60 survey participants (30.0%) perceived telemedicine services to be inferior to in-person consultations. Interview participants elaborated and expressed that telemedicine is lacking in multiple functions of care including laboratory tests and diagnostics, physical assessment, and rapport-building. Their preference for telemedicine or in-person visit depended on the health condition. A participant with an atypical presentation of her illness and needed multiple laboratory tests for her diagnosis: “I would never use telehealth consultation again for other matters besides follow-up care.” (21-25, female). She explained that the telemedicine consultation was not useful because she still needed to do an in-person consultation to have her concerns addressed. She also mentioned that whether or not she did telemedicine or an in-person consultation, she still had to be at the hospital for laboratory results.

Several respondents noted that some diseases cannot be assessed through telemedicine due to the necessity for certain equipment or physical assessment, leading to the preference for face-to-face consultations:

> “But for those illnesses that cannot be diagnosed by video call, like those that need additional equipment to check, then it’s better to do it face-to-face.” (26-30, female)
>
> “I used to have skin asthma. So for me, it’s really necessary to go and see a dermatologist so that he/she can physically see what rashes I have.” (41-45, female)

### Factors influencing telemedicine use and satisfaction

#### Facilitators

##### Safety of telemedicine during the pandemic

All 16 interviewees cited COVID-19-related reasons for their telemedicine consultations. Many participants used telemedicine because of the possibility of being exposed to the virus on the way or at the place of face-to-face consultation itself:

> “Telemedicine has less exposure [to the coronavirus], less travel time and it’s also related to my mental health wherein I really don’t want to leave the house.” (26-30, female)
>
> “With the pandemic, of course you’d choose to not expose yourself further. If you’re already sick, you don’t want to expose yourself to an additional kind of virus that’s more deadly.” (26-30, female)

##### Telemedicine offers options that maintain privacy

A number of interviewees preferred telemedicine because privacy could be maintained. Being able to be discuss their chief complaints and questions in history taking pertaining to their private areas were some of the reasons they chose telemedicine:

“Telemedicine is convenient for me. You’re still one--on--one with the doctor. For example, either I’m in the living room or in my bedroom. Pre-pandemic wise, before, in the clinic, sometimes there are other doctors who share a cubicle, especially if, let’s say, the doctor is asking something regarding your private area, sometimes you are ashamed to mention it because others might hear.”36-40, female)

> “In my case, I couldn’t go out [because] it’s a sexual concern.” (21-25, female)

##### Telemedicine is affordable due to reduced costs

Around 128 survey participants (64.0%) identified cost as the reason they chose telemedicine. Interviewees supported this and mentioned being able to save on transportations costs and that the doctor’s fees were much more affordable than before:

> “For me, I was able to save with telemedicine. Transportation wise, I didn’t need to travel] I’m not sure if it depends on the doctor’s fee, but so far it seems that the doctor’s fee is cheap and only costs around USD 7-11 per consultation.” (36-40, female)

##### Telemedicine is convenient due to reduced time and travel requirements

Most of the survey participants identified convenience as a facilitator to telemedicine use and satisfaction. Most interviewees also recognized that telemedicine is more convenient compared to face-to-face consultations for the following reasons: reduced (or absent) waiting line which additionally removes the necessity to file a leave of absence to go to the doctor, the absence of traffic, and the elimination of niceties when going outside such as taking a bath and dressing up:

> You don’t have to get dressed, and drive or take a Grab.” (16-20, female)
>
> “There are more chances that the video chat will definitely save a lot more time. There’s no travel time, there’s no waiting time.” (26-30, male)
>
> “With telemedicine generally at least inmy experience, the waiting time is reduced so I think in that regard it’s nice.” (16-20, female)

##### Telemedicine is easily accessible and readily available

Another facilitator to telemedicine use and satisfaction was accessibility in availing the services, which was identified by 160 survey participants (80.0%). The interviewees explained that telemedicine services were available at any time and did not require them to see their doctor physically:

> “Access [was one of the reasons why I chose to use telemedicine] because you just wait in the house and/or the doctor’s availability.” (51-55, male)
>
> “I like telemedicine because when you need it and you’re far from your doctor, you can just call and describe and maybe send pictures or information. You can still get your medicine and advice from the doctor.” (41-45, female)

##### Telemedicine offers more avenues of communication

Telemedicine offers more avenues for communication as its scope includes text-based messaging, voice calls, and video calls across different platforms. In the interviews, several platforms were identified including KonsultaMD, SeriousMD, and Aide. Messenger and Viber were noted by some participants to be convenient applications for communication because they can reach out anytime. Hospital hotlines and school medical services were also platforms mentioned by interviewees. This was perceived as a benefit of telemedicine in itself:

> “If you are out of the WiFi zone, it is hard to connect with video call. You have the option to email, text, or call.” (56-60, female).

Survey respondents used one or more of the following: SMS, messaging applications, email, video call, voice call, and telemedicine-specific platforms. About half have used messaging applications (50.0%) and video call (45.5%) for telemedicine. Email (11.5%) is least used for telemedicine (**Table 3**).

**Table 3.** Applications and platforms of telemedicine (n=200)

| **Telemedicine applications and platforms** | **N (%)<sup>a</sup>** |
| --- | --- |
| SMS | 54 (27.0) |
| Messaging applications (Messenger, Viber) | 100 (50.0) |
| Email | 23 (11.5) |
| Video call | 91 (45.5) |
| Voice call | 63 (31.5) |
| Telemedicine-specific platforms (e.g., SeriousMD, Konsulta MD) | 46 (23.0) |

<sup>a</sup> N does not total to 200 since each participant was permitted to select all methods of telemedicine they have used.

### Barriers

#### Perceived poor service quality due to limited to no prior patient-physician relationship

This limited to no previous physician-patient relationship results in dissatisfaction with the services because of perceptions on poor service quality:

> “That’s also the weakness of that telemedicine platform [redacted]. It’s because you’re queueing for doctors, for GP doctors, right? What happens is that you don’t get to choose. Whoever is available, that’s who you’re getting.” (21-25, female)

#### Perceived lack of experience among telemedicine providers

Depending on the telemedicine service and platform used, some interview participants were unable to choose a physician and were only able to consult with whoever was available during their telemedicine consult:

> “But for emergency cases, it’s mostly resident doctors who would answer [the telemedicine hotline], not really a doctor [consultant/attending]. I experienced that in [redacted hospital 1], when they weren’t sure if they should ask their superior, or rather the department head of dermatology, what should be done to me. This means they couldn’t make decisions on the spot about what should be done to the patient, unlike in [redacted hospital 2], decision making is automatic because it’s really a doctor answering.” (31-35, male)

#### Inherent limitations of telemedicine

A number of participants expressed concerns on service quality of telemedicine due to its limitations, especially for conditions that require diagnostic tests and physical check-ups. Doctors ask several questions and seek validation from patients. There is also perceived poorer service quality because patients feel that they are not being checked thoroughly by the doctor:

> “I don’t think the consultation can provide enough accuracy compared to an in-person consultation for the prescribing doctor. I don’t think an over-the-phone conversation can truly give her an accurate evaluation of myself.” (26-30, male)
>
> “It’s really different when the doctor looks at you, puts his stethoscope on you, feels your body you know. Unlike in the past, the doctors will immediately touch the part of your body that is painful.” (51-55, male)

#### Perceived high costs

The cost of telemedicine was perceived as a barrier to the use and satisfaction of telemedicine services with participants expecting that costs are lower. As one interviewee remarked: “I really expected it [telemedicine] to be cheaper than the physical so if I’m going to pay the same price for face to face and telemedicine, then I’ll go to the physical one since same price.” (16-20male)

In the survey, the cost of telemedicine ranged from USD 0 to USD 192 with a median of USD 7.5. Nearly half (42.5%) of the survey participants had consultations for free with some relying on promotions to avail telemedicine. There were also participants perceived the price to be expensive for others: “I was just thinking in general, how would Filipinos–from all demographics, all social classes – how would they find it? So I said it [the cost] might be a barrier for some.” (16-20, female)

#### Poor network connectivity resulting to ineffective communication

Ineffective communication as a result of poor network connectivity was identified by 5 (2.5%) survey participants as a barrier. One interview participant noted: “Even if you’re connected and you’re talking, sometimes the other person doesn’t hear what you’re saying, or vice versa. They hear you, but they don’t understand because it keeps cutting off [because of poor connectivity].” (51-55, male). Others also mentioned that their satisfaction with telemedicine depended on how smoothly the telemedicine consultation goes, which in turn is significantly influenced by internet connectivity, the platform’s data usage, and the gadgets used for the consultation: “If we’re in the middle of a serious discussion, then suddenly it [the internet connection] will cut off? It’s awkward and embarrassing, especially if I don’t know the doctor.” (41-45, female)

#### Inaccessibility of required technology interferes with telemedicine use and satisfaction

Participants cited inaccessibility to technology as a factor influencing its disuse and dissatisfaction. They identified access to technology required for the consultation to be an important consideration: “People don’t have mobile load. Some don’t have good cameras for their phones or gadgets, or some don’t have it. I think that’s the disadvantage of using telemedicine.” (41-45, female)

## DISCUSSION

Our study showed that patients are generally satisfied with the services provided through telemedicine applications and platforms. This is consistent with previous studies that report high levels of patient satisfaction (23–25). Telemedicine was perceived to be similar to in-person consultations in that the participants were able to obtain medical advice and have their health concerns addressed regardless of the mode of delivery. Some perceived it to be better primarily because of convenience and accessibility. However, the inherent limitations of telemedicine restrict its utility, especially for health conditions that require physical assessments and laboratory tests.

We found that telemedicine use and satisfaction are influenced by a number of factors including: safety during the pandemic, privacy, affordability, convenience and accessibility, and availability of more avenues of communication. Safety was a major concern that prompted participants to use telemedicine. Telemedicine enables patients to avoid situations that would expose them to SARS-CoV-2, the causative agent of COVID-19, such as traveling and staying for long periods in high-risk environments. These safety concerns, together with lockdown restrictions, resulted to significant declines in hospital admissions for non-urgent procedures (2). Innovative solutions through telemedicine have been introduced including video visits (25). Participants also mentioned that telemedicine assisted in maintaining privacy. The benefits of anonymity are especially important with regards to sensitive and potentially stigmatizing health issues such as mental or sexual health conditions (29). Because telemedicine removes the need to travel, participants also viewed it as more affordable and convenient. This was noted by participants as an enabling factor to use telemedicine, especially since a third of our participants are full-time employees, while a quarter are students. Traveling for healthcare purposes could mean missing work or school (26), and telemedicine therefore gives them greater ability to manage their time around consultations. Similarly, the variety of communication modes and platforms available contributed to patient use and satisfaction (27). This enables patients to continuously communicate with physicians should technical difficulties arise.

Meanwhile, barriers identified were perceptions on poor service quality arising from lack of prior physician-patient relationship, lack of experience, and inherent limitations; perceived unreasonable costs; and poor internet connectivity and other technological barriers (e.g., gadget availability and specification). Reduced trust in the physician can leave the patient unsatisfied with the service provided and affects patient compliance with the doctor’s advice (28). Established relationships are an important factor in telemedicine use, as patients are less willing to use telemedicine to see a provider that they do not know (29,30). While some participants in our study, as well as published literature, noted cost as a factor contributing to patient satisfaction (25,31,32), we also found cost to be a reason for dissatisfaction among our participants. This may be due to the significant proportion of participants in our sample who were not employed with almost a half not enrolled in any health insurance plan. The Implementing Rules and Regulations (IRR) of the Universal Health Care (UHC) Act stipulates that the Philippine Health Insurance Corporation or PhilHealth shall use its contracts to incentivize the integration and use of telemedicine (33). The PhilHealth Konsulta package is a comprehensive outpatient benefit that integrates telemedicine to ensure access to services (34). According to a PhilHealth circular released in 2021 (35), home isolation services including telemedicine will be incentivized as long as Konsulta providers have accomplished all necessary documents. In addition, several health maintenance organizations in the Philippines reimburse telemedicine consultations which lessens the burden on patients (36–38). This statement is supported by Polinski et al. (39), stating that medical insurance provides care at lower costs. However, because of the pandemic, the rollout of the Konsulta package has been significantly delayed and therefore, patients are unable to avail of the services at a lesser cost. In addition to issues of cost, poor network connectivity and technological barriers decrease levels of patient satisfaction (40). These barriers are especially significant in the Philippines, where service delivery and resources are inequitably distributed (41). Because the country is archipelagic, there are communities with limited access to the Internet and technology. As a result, telemedicine is not widely adopted in these resource-limited communities and these barriers need to be addressed to provide services to patients where physicians and/or specialists are few (26).

A number of limitations need to be considered when interpreting our findings. First, the results of the study are influenced by the social context and implications of the COVID-19 pandemic during the time the study was conducted. Because of this, scores provided by the participants are not indicative of telemedicine alone, but rather, indicative of patient satisfaction when using telemedicine in the context of the COVID-19 pandemic. Further, patient telemedicine satisfaction studies generally report high ratings reflective of their experiences with health care and service delivery (40). However, we addressed this issue by including a measure on preference between telemedicine and face-to-face consultation (40). A considerable proportion of participants reporting the same level of satisfaction for both modes of service delivery and a few interview participants reporting less satisfaction for telemedicine. This confirms in part that telemedicine satisfaction is high in our study because of their experience with telemedicine itself, and not only because of the general care they receive from the health system. Second, our use of convenience sampling and online data collection methods potentially excluded participants from low-resource and remote communities. Patients from these areas may have other experiences, particularly barriers, in their use of telemedicine services. While we attempted to interview participants with different backgrounds and experiences, majority of those who were willing and consented were mainly from urban areas. But we were still able to capture issues of cost and technology. Third, we asked their general experience and satisfaction to telemedicine regardless of platforms. We are therefore unable to disentangle the effect of specific telemedicine platforms on satisfaction and use. Despite these limitations, our study provides a rich source of data, building on the evidence that telemedicine can be integrated into routine care during and beyond the pandemic while offering insights into use and satisfaction through the lens of patients in a low-and-middle income country.

## CONCLUSION

This study showed that Filipino patients are generally satisfied with telemedicine services provided during the COVID-19 pandemic. Telemedicine use and satisfaction are influenced by individual, health provider and system, and external factors such as technology. Our findings also suggest that participants have varying reasons for perceiving telemedicine to be equal, inferior, or better than in-person consultations. Telemedicine was viewed as safe, efficient, and effective when technological barriers are removed. However, expectations of patients on the costs, as well as the conditions that can be addressed through telemedicine, need to be managed by providers to increase satisfaction. Continued adoption of telemedicine will require improvements in technology and better patient communication related to their telemedicine provider and the associated costs. Our study points to the following recommendations: (a) Integration of telemedicine services in geographically remote areas that lack access to medical services; (b) Strengthening of infrastructure to allow the use of devices and Internet; (c) Training and performance evaluation of telehealth providers to ensure quality telemedicine services; (d) Patient communication on telemedicine and its limitations; (e) Patient support for those with technological difficulties; and (f) Future research to include stakeholder perspectives and patient experiences from remote communities.

## Data Availability

All data produced in the present study are available upon reasonable request to the authors

## Declaration of Conflicting Interests

The authors declared no potential conflicts of interest with respectto the research, authorship, and/or publication of this article.

## Funding

The authors received financial support for this research from the Ateneo Center of Research and Innovation (ACRI). The funder had no role in study design, data collection and analysis, decision to publish, or preparation of the manuscript.

## Notes

### Competing Interest Statement

The authors have declared no competing interest.

### Funding Statement

This study was funded by Ateneo Center of Research and Innovation (ACRI).

### Author Declarations

This research was given ethical approval by the Ateneo University Research Ethics Committee (SMPH 2021 Group 15).

